# Measures of Behavior and Life Dynamics from Commonly Available GPS Data (DPLocate): Algorithm Development and Validation

**DOI:** 10.1101/2022.07.05.22277276

**Authors:** Habiballah Rahimi-Eichi, Garth Coombs, Jukka-Pekka Onnela, Justin T. Baker, Randy L. Buckner

## Abstract

Locations of people moving about their lives are now commonly tracked through smartphones and wearable devices that access the Global Positioning System (GPS). Immediate measures include the estimated locations that identify visited map points and the travel paths between them. Here we introduce DPLocate, an open-source GPS data analysis pipeline designed to derive measures that abstract away from the original locations (and hence the identity of the individuals) and capture dynamics related to social, vocational, sleep, and clinical behaviors. We divide derived measures into primary and secondary. Primarily derived measures stay close to the original location data and extract deidentified metrics, including distance traveled, time spent at the main locations, and estimates of travel activity (entropy). Secondary derived measures estimate life patterns that are captured incidentally by extracting returns to the Points of Interest (POIs) in behaviorally-relevant time-bands. For example, measures of behavioral dynamics and social interactions can be gleaned by estimating the time spent in POIs across day, evening, night, and late-night time-bands. The utility of these derived measures for research is illustrated in college students and for clinical monitoring in individuals living with psychiatric disorders. Captured dynamics included behavioral transitions at the onset of the Covid-19 lockdown. Limitations of derived data are discussed, including the necessity to protect derived data from identification and possible ways in which the derived data might be misinterpreted.

---

The widespread availability of smartphones and wearable devices accessing the Global Positioning System (GPS) offers the opportunity to track individuals as they go about their lives. Recent studies suggest that the monitoring and classification of individuals’ locations during daily routines provide accessible measures of human behavior. In addition to extracting estimates of mobility, entropy, and trajectories from continuously recorded data [1,2], significant points of interest (POIs) enable researchers to investigate the relocation patterns as a window into an individual’s life [3-6]. Therefore, there is an ongoing need for open-source, secure pipelines to analyze GPS data and extract behavioral patterns on-site without exposing identifiable location coordinates.

Existing algorithms to analyze GPS data mostly rely on a combination of temporal, spatial, and conceptual features to extract the behavioral components [7]. Meanwhile, the technical and idiosyncratic characteristics of analyzing GPS data make it an evolving field of interest for investigators. Some of those characteristics include: (1) the identifiability of the GPS coordinates makes sharing or even viewing the raw data challenging [8]; (2) GPS signal loss occurs due to intermittent satellite signals, operation system block, and manual settings to avoid battery drainage [9]; and (3) the inaccuracy of the recorded GPS using different applications can bias clustering and behavioral inferences.

In this paper, extending from [10], an open-source GPS analysis pipeline called Deep Phenotyping of Location (DPLocate) is introduced. DPLocate securely reads encrypted GPS data and estimates daily, weekly, and Time-Band restricted patterns to infer sleep, work and social behaviors in addition to major life events [11]. A digital phenotyping research platform, Beiwe [12], installed on smartphones, provided data for evaluation of the DPLocate pipeline, though the pipeline is general and can build from any platform that extracts temporally-extended raw GPS data. Within DPLocate, epoch-based temporal filtering accounts for the intermittency of the GPS signal by focusing on temporally adjacent data point sequences, called Epochs, as data units instead of single data points and, consequently, mitigates the accuracy challenge by removing outliers and Epochs with insufficient data points. After applying a density-based spatial clustering algorithm to the Epochs to detect POIs, Markov Model diagrams are utilized to represent the probability of transitioning from one POI to another during behavioral-relevant Time-Bands. Time-Bands separate Day, Evening, Night, and LateNight time periods. These Time-Bands were chosen to roughly correspond to periods during which individuals work or attend school, socialize, and sleep.

The DPLocate pipeline’s output for each participant includes an abstracted GPS map with clustering results, a color-coded daily behavioral map displaying visited POIs, and Markov Model transition diagrams between POIs during each Time-Band. These graphical outputs provide a unique behavioral view of the individual that can be compared with other participants. As proof-of-concept for DPLocate, the behavioral patterns of individuals in two cohorts were tracked, along with self-reported mood measures.

## Materials and Methods

### Participants

#### Two separate samples of individuals contributed data

##### Study 1: Undergraduate Study

Four undergraduate participants (age: mean (SD) 19.8 (0.5), range: 19-20 years old; all females) were recruited from a local private institution. The study duration consisted of one academic semester, including buffers into winter and summer breaks on either side to account for natural transition periods (maximum 28 weeks). Participants were compensated as follows: per hour for the in-lab and online personality testing sessions, a milestone bonus for completing the study, and a monthly bonus to encourage continued participation. Participants were required to be enrolled full-time in classes and own a smartphone compatible with the study smartphone application, Beiwe [12]. Participants were not excluded for current or past psychiatric treatment or if they began treatment or medication for mental health issues during the course of the study. All study procedures were approved by the Institutional Review Board of Harvard University.

##### Study 2: Hospital Study

Five individuals (age: mean (SD) 30.6 (10.0), range: 24-48 years old; 1 female) were recruited from an ongoing cohort following the clinical progression of severe mental illness at a local hospital. The clinical examples in the previous actigraphy-based sleep study were selected from the same cohort [10]. Individuals were diagnosed with psychotic disorders (bipolar, n=2; schizophrenia, n=3) using the structured clinical interview for DSM-IV [13]. Participant enrollment for this study targeted obtaining >1 year of data for each participant (duration: mean (SD) 357.8 (97.9) days, range 219-495 days), which included a nearly continuous collection of smartphone data from each participant. Participants were compensated for complying with the data collection, as well as for monthly in-person study visits during which clinical assessments were recorded to quantify disease progression using clinical gold standard measures. Milestone bonuses were also provided to encourage continued participation. All study procedures were approved by the Institutional Review Board of Partners Healthcare.

### GPS Data Acquisition

Participants installed a research smartphone application, Beiwe [12,14], designed to actively and passively collect data. Beiwe has been utilized in several clinical [12,15] and behavioral studies [10,14,16-18] with different settings, and the design has been improved based on user preference and study requirements. In addition to active self-reported data collection, Beiwe also passively collects data from participants’ smartphones. In order to put less strain on participants’ phone batteries, GPS sampling was set to collect for 2 minutes every 10 minutes. However, the actual (obtained) sampling is not static and can vary based on the participant’s movement or availability of the GPS signal. As a result, some data points are recorded for a shorter time and with fewer or more gaps in between. The goal of this paper is to introduce the DPLocate pipeline, to investigate the behavior and life dynamics of individuals based on analyzing their GPS data collected intermittently by smartphone. The modules of the pipeline are explained below and shown in the block diagram in Figure 1.

**Figure 1.**
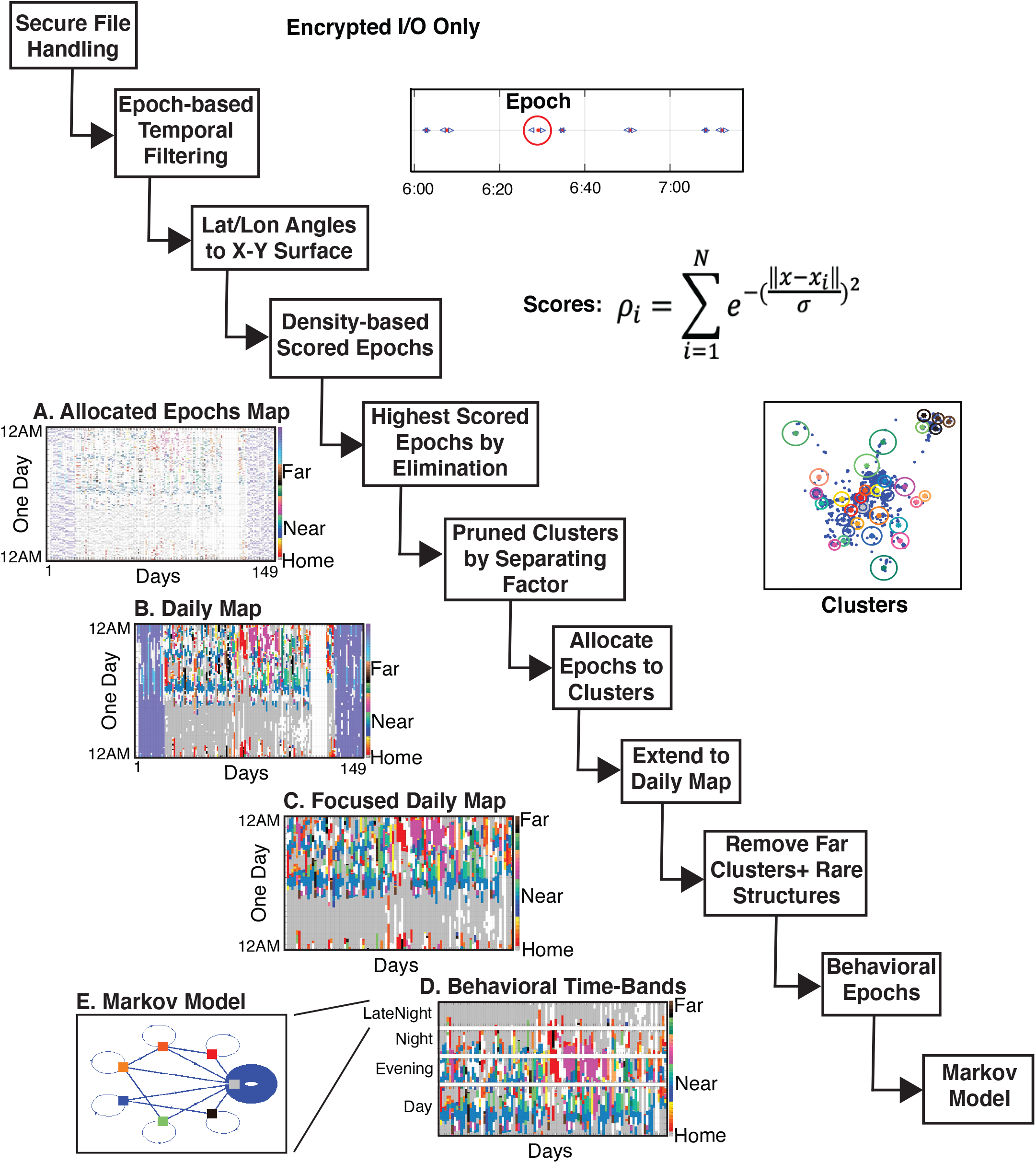
The DPLocate GPS processing pipeline. The sequential steps to process the GPS data are illustrated with a representative example. The raw and intermediate results are stored in an encrypted format to ensure secure file handling. Epochs of temporally adjacent data points that are at least 45 seconds apart from neighboring Epochs are filtered and saved. After converting the latitude and longitude angles to a standard X-Y surface, density-based spatial clustering is applied to the Epochs to find points of interest (POIs). Following the “Scores” formula, the score for each Epoch is the summation of the points in the 50 meters neighborhood of the center, calculated based on their closeness to the center. To detect POIs, the locations of the 80 highest scored Epochs are chosen by eliminating the previous Epochs and their 50 meters neighborhood. All epochs are then allocated to the detected POIs and color-coded from Near to Far referenced to the estimated Home POI resulting in the initial color-coded **Allocated Epochs Map** (**A**). Then the Allocated Epochs are expanded up to 12 minutes before and after their original time, limited to the previous and next Epochs, to create the **Daily Map** (**B**). In the Daily Map, dark gray is Home (the most visited POI), light gray represents the Epochs that do not belong to any POI, white is missing data, and the remaining colors represent distinct POIs. Because the focus of the DPLocate pipeline is to find repeating patterns, rare (less than twice occurring) POIs, relatively far POIs, and days with peculiar structure (based on the day-to-day correlation of the staying-at-Home pattern) are removed to produce the **Focused Daily Map** (**C**). The time of the day is classified into four **Behavioral Time-Bands** (**D**): Day (9AM-5PM), Evening (6PM-10PM), Night (10PM-2AM), and LateNight (2AM-6AM). Finally, a **Markov Model** diagram (**E**) is utilized to provide a visualization of the relocation behavior of the participant in each Time-Band.

### Secure Handling of the Data

Secure handling of GPS data is critical [19-21] because the GPS data files contain location positions with their recorded times, exposing identifiable information such as home, work, and even partner’s addresses. Therefore, to protect the data, all raw data are stored as encrypted files. Moreover, a systematic practice is maintained to securely delete any decrypted files generated by the pipeline in a fraction of a second post-decryption; the decrypted data are kept only in the Random-Access-Memory (RAM) of the system during the entire procedure without being stored anywhere. The result of each milestone is saved as an encrypted file under the same protected directory for the individual. Within our local implementation, the DPLocate pipeline runs behind institutional firewalls designed to secure human research data, where encrypted files are also stored.

### Epoch-based Preprocessing

The DPLocate pipeline aims to extract patterns of behaviors and markers from the semi-continuous passive data. For GPS data analysis, several studies [22-24] suggest that the POIs, the places most visited by the individual, are key anchors to analyze location-related behaviors. Accordingly, instead of tracking the participants’ trajectories, the main target of this pipeline is a discrete representation of the individuals’ daily mobility patterns based on detected POIs. Some studies incorporate their POI detection algorithm with the data acquisition procedure [23]. Other studies use semantic and adaptive methods to adjust the sampling rate [25], and still others detect the POIs based on the mobility speed [26-28]. Here we deployed a data-driven clustering approach of the raw data, which was collected on a semi-continuous basis limited to the technical availability of the GPS signal and the phones’ battery life. The pipeline’s approach to the spontaneity and discontinuity of GPS data [29-30] is to apply several filtering techniques at the preprocessing stage to increase the confidence level of location detection using Epoch-based analysis.

An Epoch is defined as a collection of 1 to 100 data points that are obtained continuously and are at least 45 seconds apart from the neighboring Epochs. In rare cases where Epochs longer than 5 minutes are detected, they are broken into smaller Epochs less than 2 minutes long. A table of the parameters of the preprocessed data is formed with the information of each Epoch in every row that are used in preprocessing step (Table I). As reported in the literature [3,31,32] and confirmed by our collected data, the smartphones sometimes lose the GPS satellite signal and rely on the less accurate cell-tower estimated coordinates, which yields jitter in the recorded data [4]. Therefore, only the Epochs with more than 10 data points (*num_loc*) are included so that coherence of data points could be checked using statistical analysis. Additional filtering is then applied to recalculate the center of the Epoch (*mean_loc_lat, mean_loc_lon*) by excluding extreme points outside a 3 standard deviation distance from the Epoch’s original center (*num_loc_3sd*). The Epoch-based analysis acts as a filter to remove artifacts and unclear transition points in the GPS signal. Each Epoch, regardless of its number of raw data points, is counted as a single point represented by its geographical center (*mean_loc_lat_filt, mean_loc_lon_filt*) for later analyses.

**Table I:**
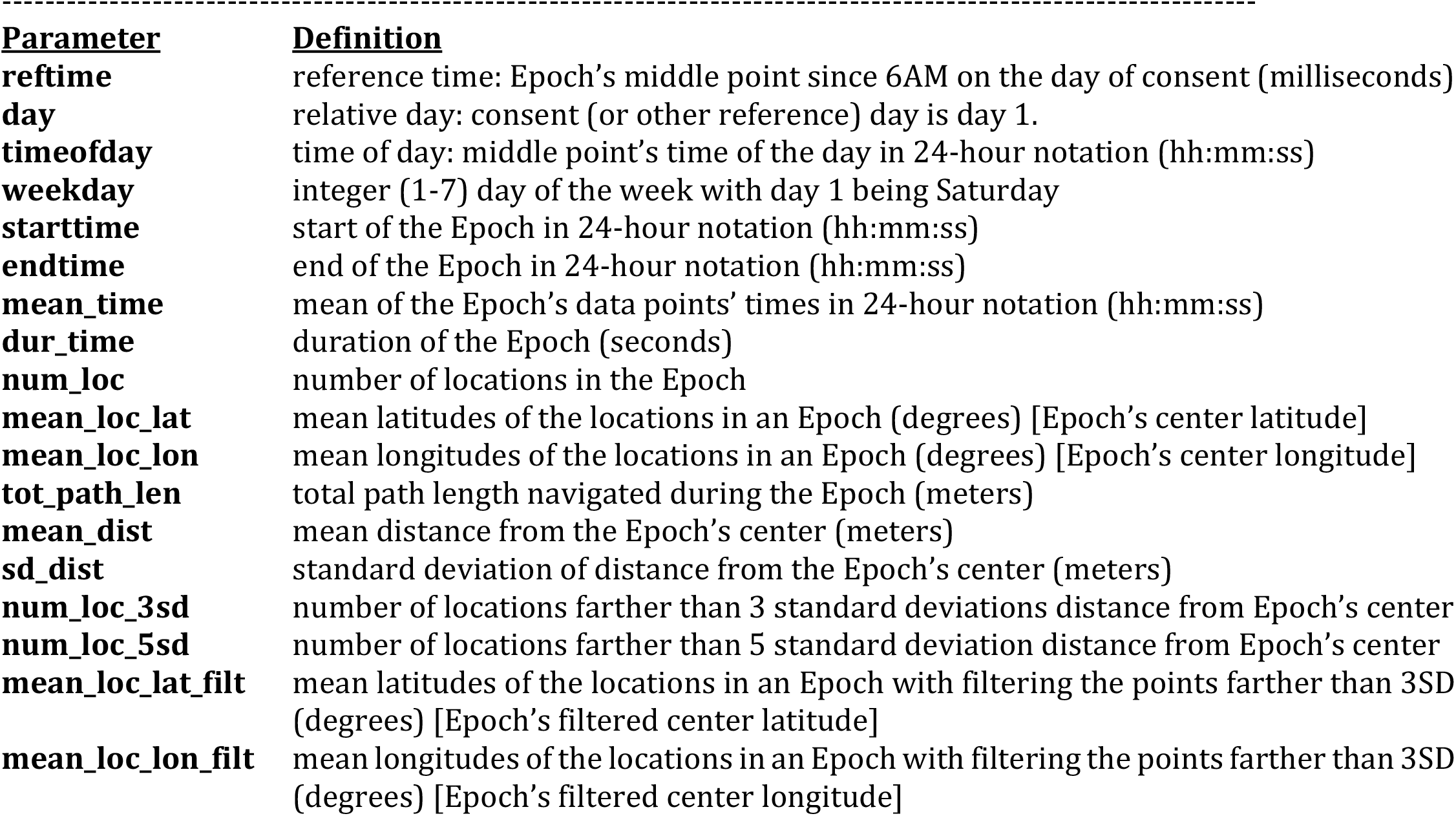
The Epoch’s parameters’ descriptions

### Point of Interest (POI) Detection Algorithm

After preprocessing the GPS data using the Epoch-based filtering, the goal of the DPLocate pipeline is to detect the visited POIs. Extensive research has been conducted to design and utilize various versions of POI detection algorithms for different applications [3,22]. Multiple approaches base their clustering on the spatial distribution of the collected data [33,34]. Accordingly, DPLocate deploys a spatial density-based clustering algorithm, which is a modified version of the Density-Based Clustering Algorithm with Noise (DBSCAN) [35,36], to find the 80 most visited locations of the individual during the data collection period. Subsequently, the algorithm then refines the locations to the 80 locations with the maximum number of Epochs in their 50 meters neighborhood weighted by their proximity to the center. After converting the geographical coordinates into an x-y spatial map, the score for each location is calculated based on the equation shown in Figure 1 (i.e., the sum of the inverse exponential distance of all points in the 50 meters neighborhood). POIs are chosen sequentially by excluding the data points in the 50 meters neighborhood of the already selected POIs.

To ensure the adequate separation of the neighboring areas for all POIs, in the first round we select the 150 most scored points; in the second round, a separation factor of 85% is applied between clusters. The points with more than 15% (one minus 85%) data in common with each other are combined to make one point. The 80 highest scored points are then selected as the POIs of the individual. An example of these detected POIs, represented by their centers and 50 meters neighboring circles, are demonstrated on a geographical map to show their distribution and distance from each other (Figure 2).

**Figure 2.**
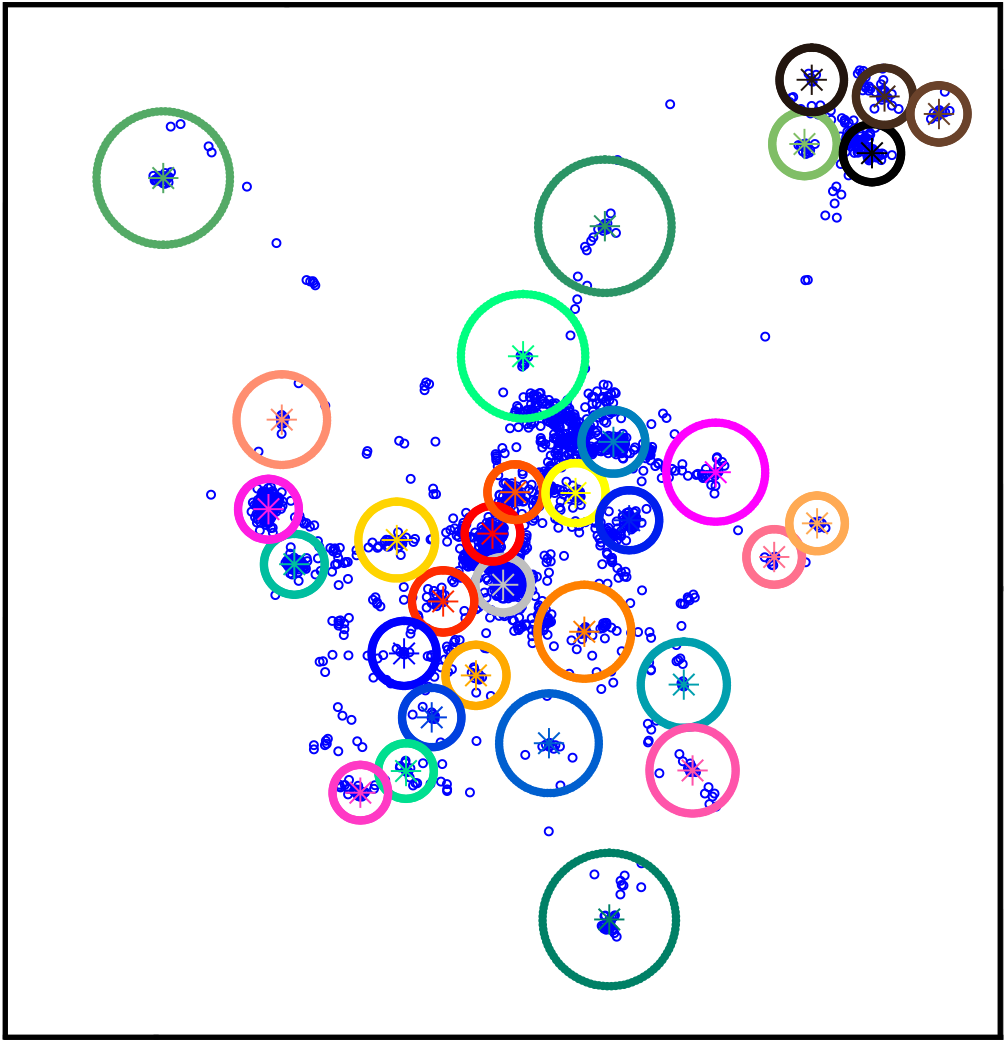
An Example of Spatially Clustered Epochs. Mean coordinates of the Epochs (small blue dots) are demonstrated on an X-Y surface for a representative example. The large colorful circles show the results of density-based clustering with the stars at the center of the circles showing the POIs and the areas of the circles as the coverage areas of the POIs (i.e., all the Epochs in the circled area belong to the POI cluster). The gray circle is called Home as the most visited location. The actual data points have been shifted and relocated in this figure to avoid identifiability.

### Assigning Epochs to the POIs

After identifying the POIs, the next step assigns each Epoch to one of the selected POIs. Since DPLocate is designed to investigate the behavioral dynamics of the individual rather than to pinpoint their exact locations, the coverage area of each POI is defined as a circle centered at the POI with a radius of 120 meters bounded by the neighboring POIs. If the point is in the coverage area of one or more POIs, the Epoch is assigned to the closest POI. Visualization of the assigned Epochs is a valuable tool to investigate the individuals’ behavior before any attempt to extract mathematical models. Consequently, based on the Epoch’s time and the assigned POIs, a color-coded daily map, as shown in Figure 3A, is produced for each individual with the x-axis demonstrating the relative day numbers (started at the consent day in our studies) and the y-axis showing the daily hours from 12AM to 12AM. The x-axes in the daily maps in this paper are shuffled to further protect the identity of the individuals. In these daily maps, dark gray represents the estimated Home location, the most visited POI. In the case where the Epoch is not in the coverage area of any POI, it is considered as unknown location shown with light gray color; otherwise, the allocated Epochs are colored based on the clustered POI colors on the geographical map. In the final step, the assigned colors are extended to 12 minutes before and after the Epochs’ times, limited to half the distance from the neighboring Epochs (Figure 3B).

**Figure 3.**
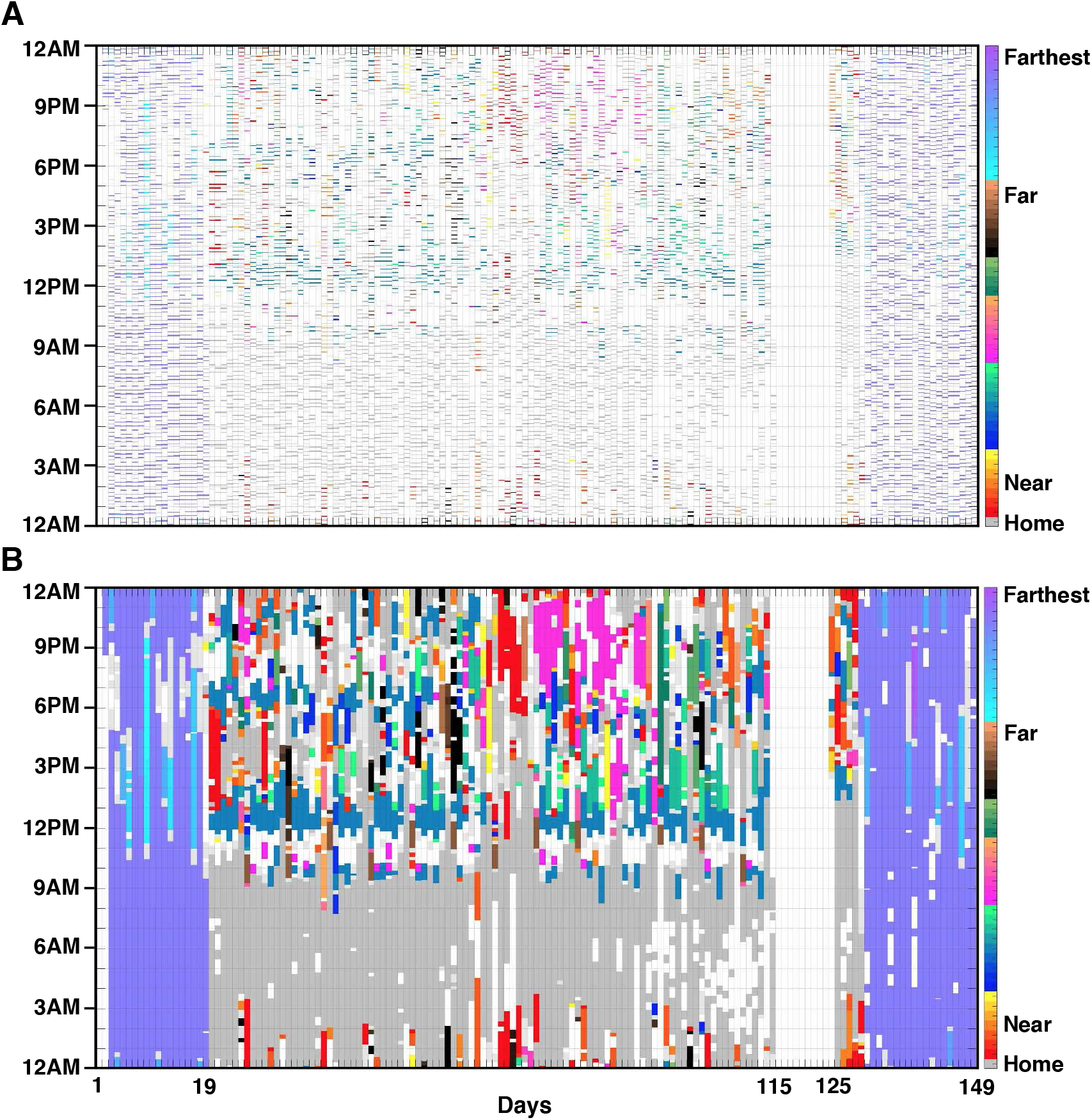
Color-Coded Daily Map. After the POIs are identified using the density-based spatial clustering algorithm, the sequential Epochs are allocated to their closest POIs if they are in their coverage areas. Example maps are illustrated for a representative participant. The x-axis is shuffled to protect the identity of the individual. **Allocated Epochs Map (A)**. The epoch assignments are illustrated in a color-coded format that displays one whole day in each column (y-axis) and the sequential days along the x-axis. Time is depicted as 24 hours from 12AM to 12AM with bottom-up direction on the y-axis. Every minute that has an Epoch of GPS data is displayed with the corresponding POI color, which can be dark gray (Home) or other colors based on the distance from Home (color-coded Neat to Far). The minute with available GPS data but not allocated to any POI are colored in light gray, and the white indicates no available data. **Daily Map (B)**. The epochs are expanded to 12 minutes before and after, limited to the half distance of the neighboring Epochs.

### Markov Model

The color-coded daily map of locations is a powerful, intuitive presentation of the individual’s daily routines during life episodes such as school, work, and weekends. Several prior studies have sought to extract mathematical models such as enhanced versions of Markov [37-39], Bayesian [40], and Dynamic Network [41] models in order to predict the next location of the individual based on the previous locations. In DPLocate, however, the focus is on extracting and modeling the daily patterns and recognizing various behavioral phenotypes rather than an accurate prediction of the individuals’ upcoming change in location. Therefore, Markov Model diagrams are deployed with some data modifications and adjustments to demonstrate the stationary repeating patterns. As a preprocessing procedure, the POIs farther than 25 kilometers from Home, and POIs that happen only once during the study are removed from the map. Further, the pipeline correlates staying at the Home location during each day with other days, and calculates a similarity parameter using a moving window to detect and remove the days with rare structure, such as nights that occur at the end of the semester or travel days in our studies (Figure 4A). As a final preprocessing step, interpolation is used to fill in the brief missing periods with the nearest location, excluding any hours with fully missing data (Figure 4B). These preprocessing steps focus on the more stationary behaviors of the individual at the expense of losing some rare behavioral events. However, the users can choose not to exclude those rare events by minor modifications to the code. An example of the daily map with the missing days removed and the x-axis modified based on the existing days to illustrate the behavioral pattern is presented in Figure 5.

**Figure 4.**
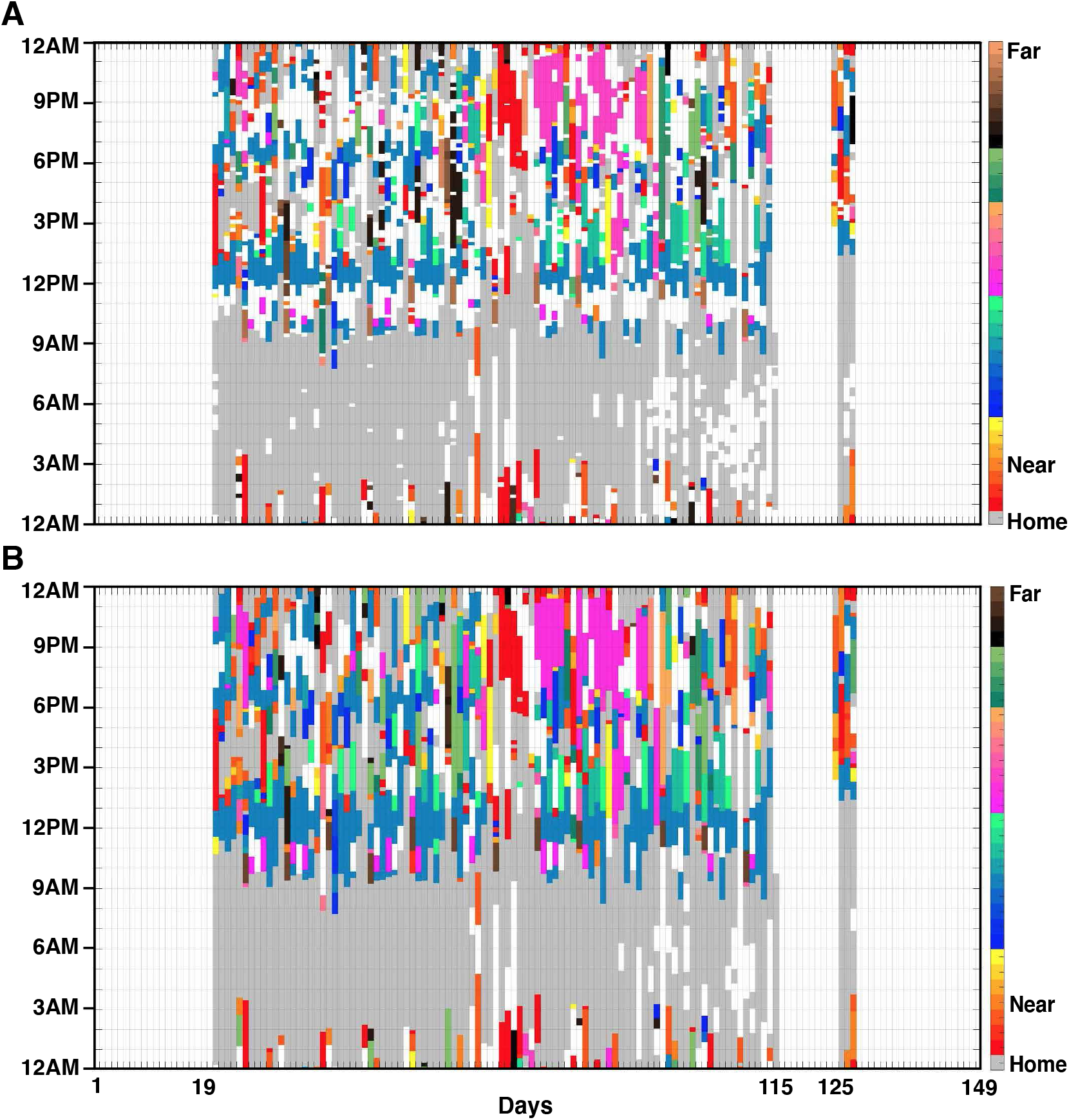
Removing Far and Rare Locations. This figure and the next illustrate the process of preparing the Daily Map to model the repeating behavior using the Markov Model. We remove the far POIs, which are farther than 25 kilometers in this case; rare POIs, which are occurring less than twice during the study; and rare structures, identified based on the day-to-day correlation of staying at Home **(A)**. The remaining allocated POIs are then extended to their nearest neighbors to cover the times where there is missing GPS signal yielding the **(B)**.

**Figure 5.**
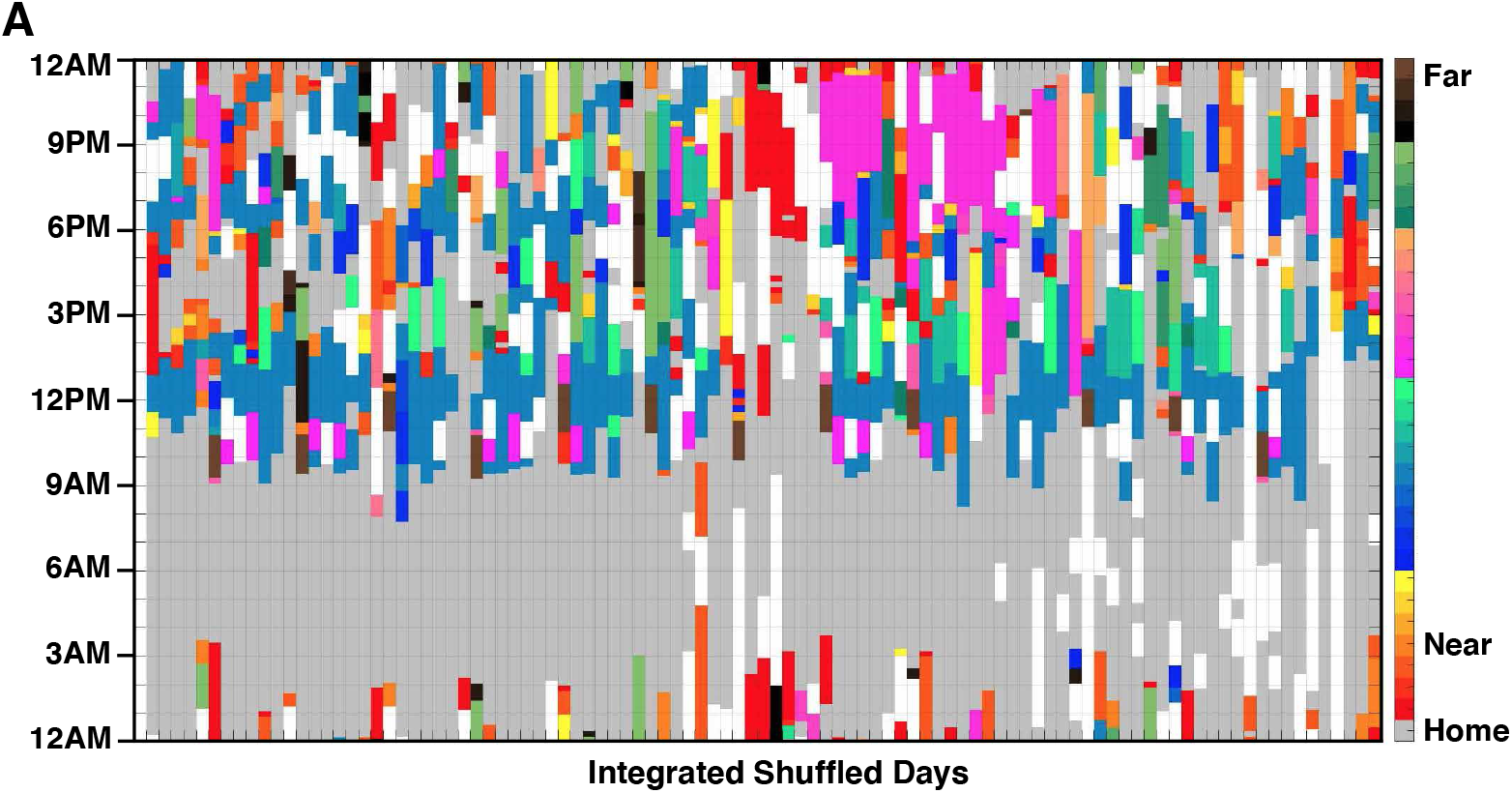
Focused Daily Map. The color-coded map after removing the rare-far POIs is replotted by removing the days with no data. As a result, the number of integrated days in this figure is smaller than the number of study days.

Another assumption incorporated into the pipeline to estimate the Markov Model transition diagram is partitioning the day into four Time-Bands, including Day (9AM to 5PM), Evening (6PM to 10PM), Night (10PM to 2AM), and LateNight (2AM to 6AM) for each individual as shown in Figure 6A. Time-Band epochs are based on the usual activity time of typical individuals but are not tailored to account for atypical life patterns (e.g. a night worker). The gaps between 5PM to 6PM or 6AM to 9AM are left intentionally to keep the Time-Band epochs associated with the regular activity time of typical individuals. Applying all these adjustments, the Markov model represents the probability of moving from one POI to another in the next 15 minutes during a specific Time-Band. The transition probability matrix for the Markov model is calculated separately for each individual without using the group data to build the transition model [42,36]. Based on the calculated probabilities, transition diagrams as shown in Figure 6B are depicted for each Time-Band, where the POIs are the nodes with corresponding colors, and the thickness of the edges represents the probability of changing location to another POI. The thickness of self-repeating edges reflects the relative staying time at that POI during the Time-Band.

**Figure 6.**
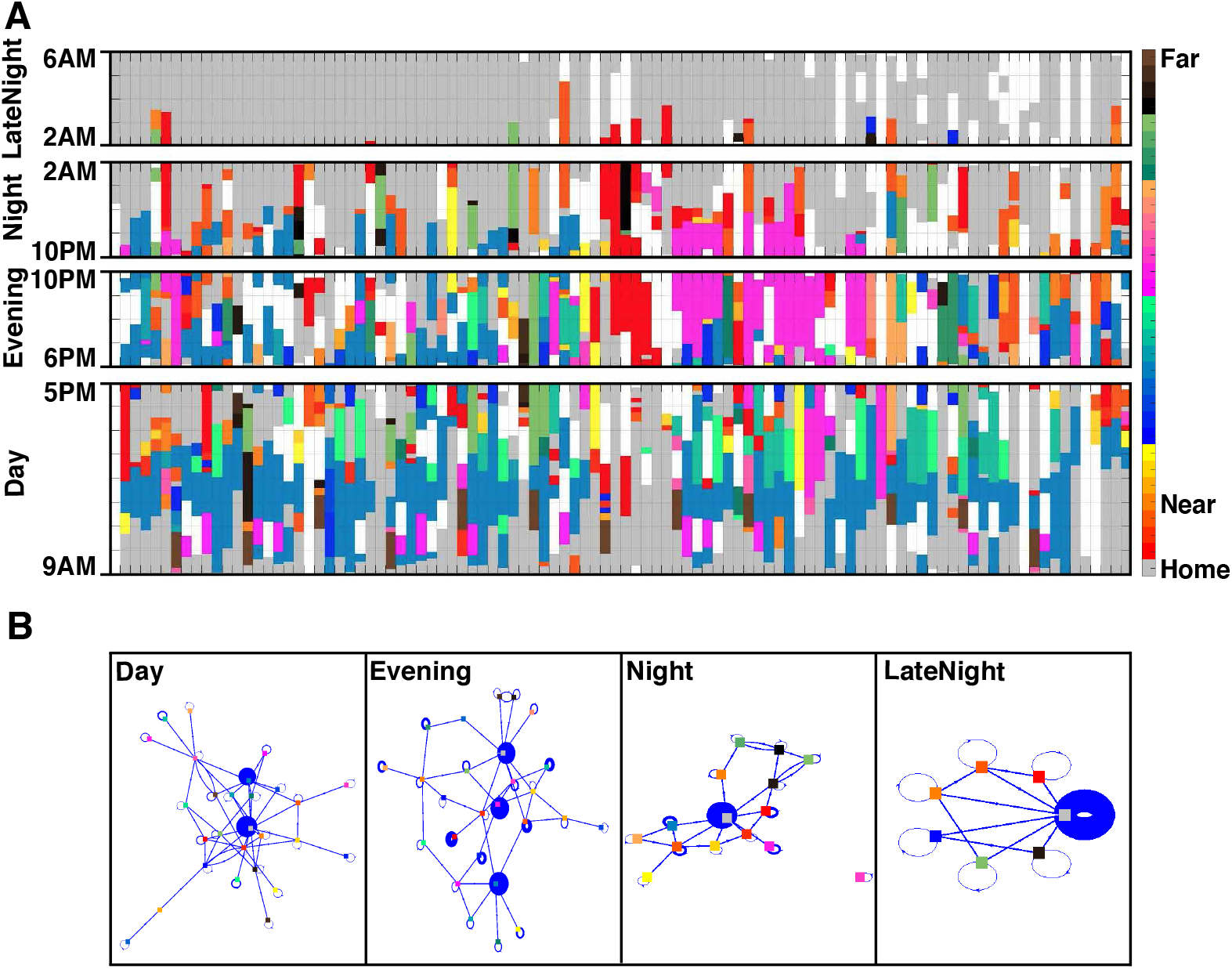
Behavioral Time-Bands and Markov Models for an Individual with Multiple LateNight Visit Locations. The hours of the day are divided into four Time-Bands Day(9AM-5PM), Evening (6PM-10PM), Night (10PM-2AM), and LateNight (2AM-6AM) chosen based on a typical schedule. The Focused Daily Map is illustrated separated into the four Behavioral Time-Bands **(A)**. The gaps between 5PM to 6PM or 6AM to 9AM are intentional to separate Time-Band epochs. Then, Markov Model diagrams **(B)** are used to demonstrate the transitions between POIs. Each panel shows the Markov Model diagram with the POIs as colorful nodes, and the probability of transition between POIs as the thickness of the blue edges. This example illustrates an individual with a busy Night and LateNight schedule, with multiple stays away from the Home location until past 2AM. Even though Home is the dominant location, there are six different POIs in the LateNight diagram, all of which eventually lead to Home.

### Code availability

The DPLocate pipeline software package is available [11] as an open-source package to be downloaded and used by the research community.

## Results

### Behavioral Phenotyping Based on LateNight Behavioral Time-Band Activity Model

The DPLocate pipeline (see Methods, Figure 1) is developed to investigate the relocating behavior of the individuals using GPS data. The main outcome of this pipeline is the patterns of relocations during Behavioral Time-Bands. A common phenotype that emerged was identifying individuals with only one location during the LateNight Time-Band. This phenotype, as an example (P2), is presented in Figure 7, shows the all-gray LateNight Time-Band map with a single-node Markov Model diagram. An observation in this example is that the individual also has a simple Night Time-Band activity pattern regardless of their Day Time-Band activity pattern. Another phenotype that emerges is some individuals display 2 or more distinct LateNight locations without changing place during this Time-Band. A relevant example (P3) is presented in Figure 8. The Markov Model diagram of this participant is represented by two separate nodes with rare transitions between them and a high probability of staying at the current LateNight location overnight. The frequency and randomness of staying at the other-than-Home location support the possibility the individual is visiting a partner’s place. Another observation out of this phenotype is that the individual spends the Night Time-Band at the same place where they will stay later that night.

**Figure 7.**
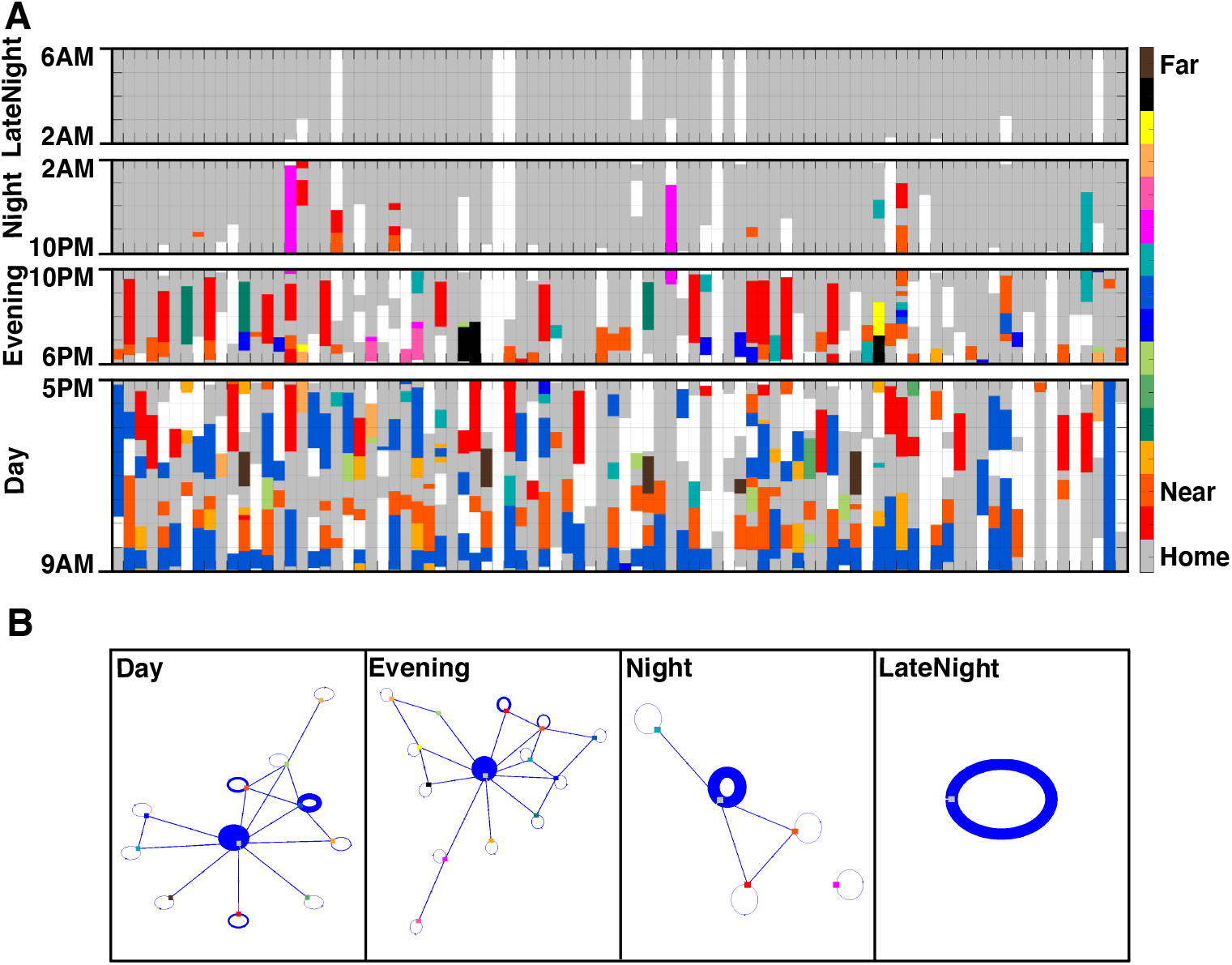
Behavioral Time-Bands and Markov Models for an Individual with a Single LateNight Visit Location. Similar to Figure 6, the Behavioral Time-Bands **(A)** and Markov Model diagrams **(B)** are shown for an individual with a single LateNight visit location. This individual shows the pattern of staying Home between 2AM and 6AM every night. The Markov Model diagram only has one node (Home) with a certainty of always staying Home. Similarly, the Night Time-Band and its associated Markov Model for this individual reflect a mostly stay-at-Home pattern with relatively few visited places.

**Figure 8.**
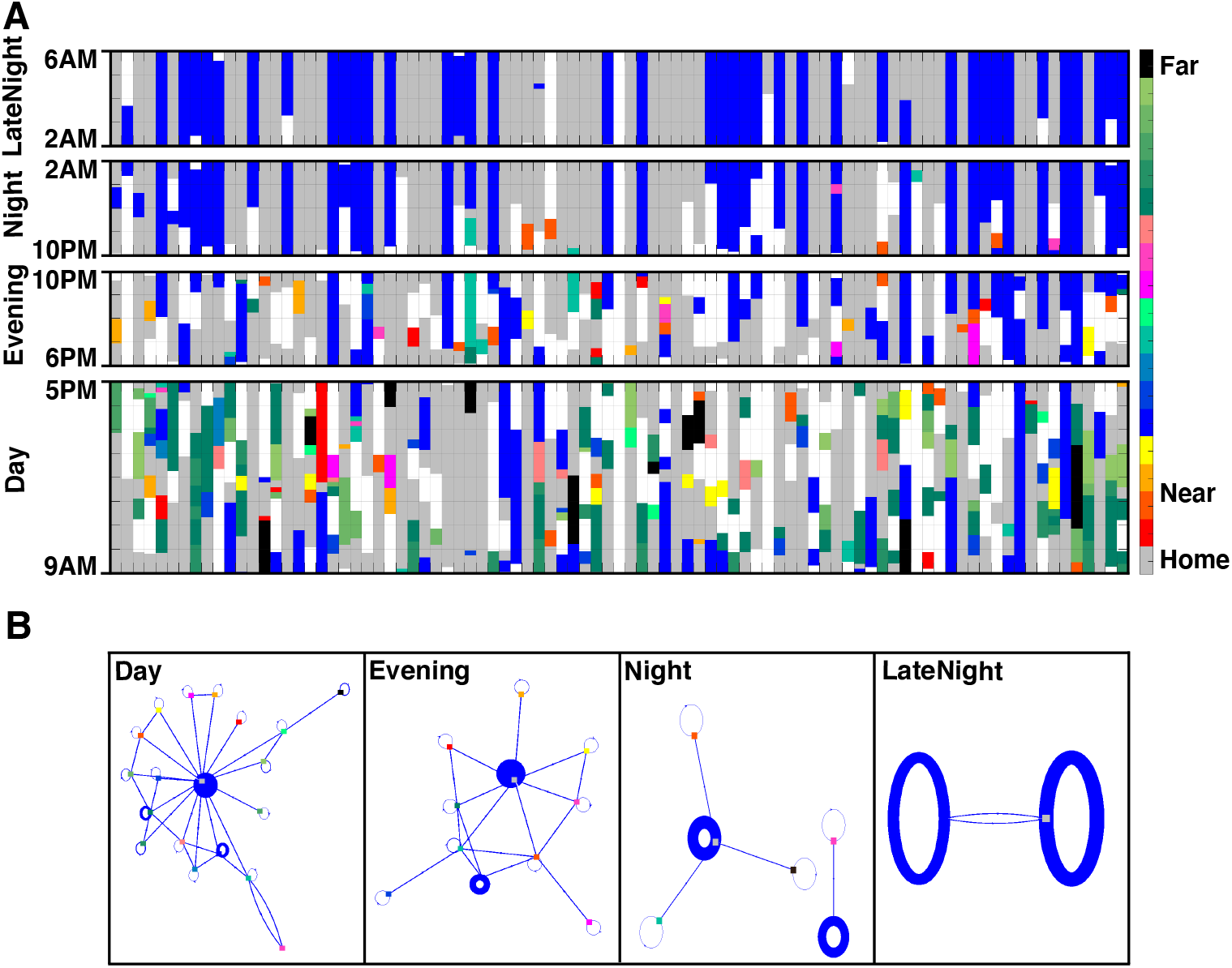
Behavioral Time-Bands and Markov Models for an Individual with Two LateNight Visit Locations. Similar to Figure 6, the Behavioral Time-Bands **(A)** and Markov Model diagrams **(B)** are shown for an individual with dual LateNight visit locations. The LateNight locations of the individual alternate between Home (gray) and another location (blue). With few exceptions, the individual rarely transitions between the two locations in the same night. The Markov Model diagram of the LateNight Time-Band represents this behavior as two thick circles with a weak connection. Moreover, the Night and even part of the Evening Time-Band, each have two dominant nodes in their Markov Model diagrams, reflecting the separation between the visited locations.

Another phenotype that is common among the college students is staying at various locations before going back Home late into the night. An example (P1) of this phenotype was presented in Figure 6 of the Methods section, where the Markov Model diagram shows staying late at different locations other than Home before eventually going back Home at the end of the LateNight Time-Band. The locations visited during LateNight Time-Band, are close but still different from Home. We cannot estimate with certainty when all of these individuals are sleeping unless we have the wrist-band accelerometer data that were comprehensively analyzed in [10]. However, the location change during the LateNight Time-Band suggests that sleep happens much later into the night. Later analyses (Figure 11) show that if the LateNight Time-Band is shifted forward for two hours for this individual, one sees the same always-at-Home location pattern as seen for P2. Moreover, the Night Time-Band of this individual is expectedly busier compared to the other previous examples. In addition to the places visited during the LateNight Time-Band, around 10 other locations are visited rather frequently, with numerous switching among them during the Night Time-Band.

### Disturbed Sleep is Detected in An Individual with Severe Mental Illness

DPLocate produces GPS-related color-coded daily behavior maps that reveal idiosyncratic sleep and life patterns that may be clinically relevant. For example, Figure 9 shows a behavioral map of an individual (P4) with a disturbed sleep pattern. The color-coded daily map during the LateNight Time-Band shows again that multiple locations have been visited during this Time-Band. The striking observation about this individual is the occurrence of leaving Home in the middle of the night, visiting several places of interest and returning back Home after 15 minutes to 1 hour trips. The disturbed sleep also includes occasions of leaving Home earlier during the Night Time-Band (after 12AM) and not coming back Home until the LateNight Time-Band (before 3AM). As expected, the Markov Model diagram of this participant during the LateNight Time-Band is noticeably busy with several places that the individual switches among, as well as several back-and-forth movements from and toward Home. Later analyses show that, unlike the previous example (P1), shifting the Time-Band forward does not find an always-at-Home epoch for this individual.

**Figure 9.**
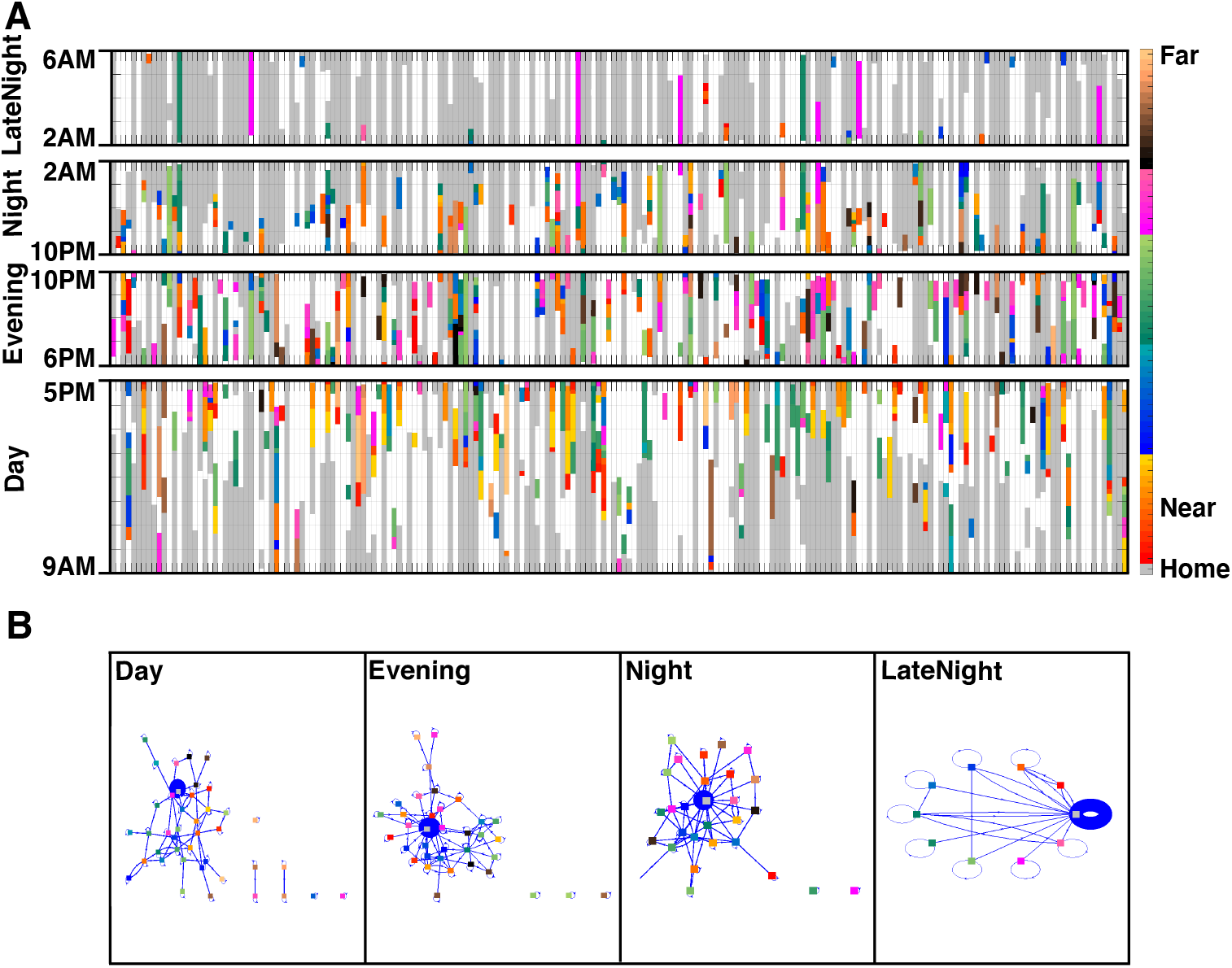
Behavioral Time-Bands and Markov Models for an Individual with Disrupted Sleep. Behavioral Time-Bands **(A)** and Markov Model diagrams **(B)** of an individual with a disrupted LateNight schedule are depicted. The thickness of the days (columns) is reduced compared to prior diagrams, reflecting the extended duration of the recorded data. The Daily Map shows sporadically occurring time outside the Home location during the LateNight Time-Band where the individual leaves Home sometime after 2AM, stays at different POI location for about an hour or more, and then goes back Home. The busy Markov Model diagram for LateNight and Night Time-Bands capture this behavior, which may be an indicator of disrupted sleep for this individual living with mental illness.

### Life Event Detection

In addition to sleep pattern phenotypes, the GPS analysis with DPLocate detects remarkable events during individuals’ lives. As an example, the case (P5) presented in Figure 10 shows two similar events during the study where the daily routines are interrupted each time for about a week. The participant stays at the same location, not very far from Home, almost all day and night. The color-coded daily behavioral map shows that some other points which are close to the same location have been visited frequently during other days, and the medical records confirm that the individual has been hospitalized during that time.

**Figure 10.**
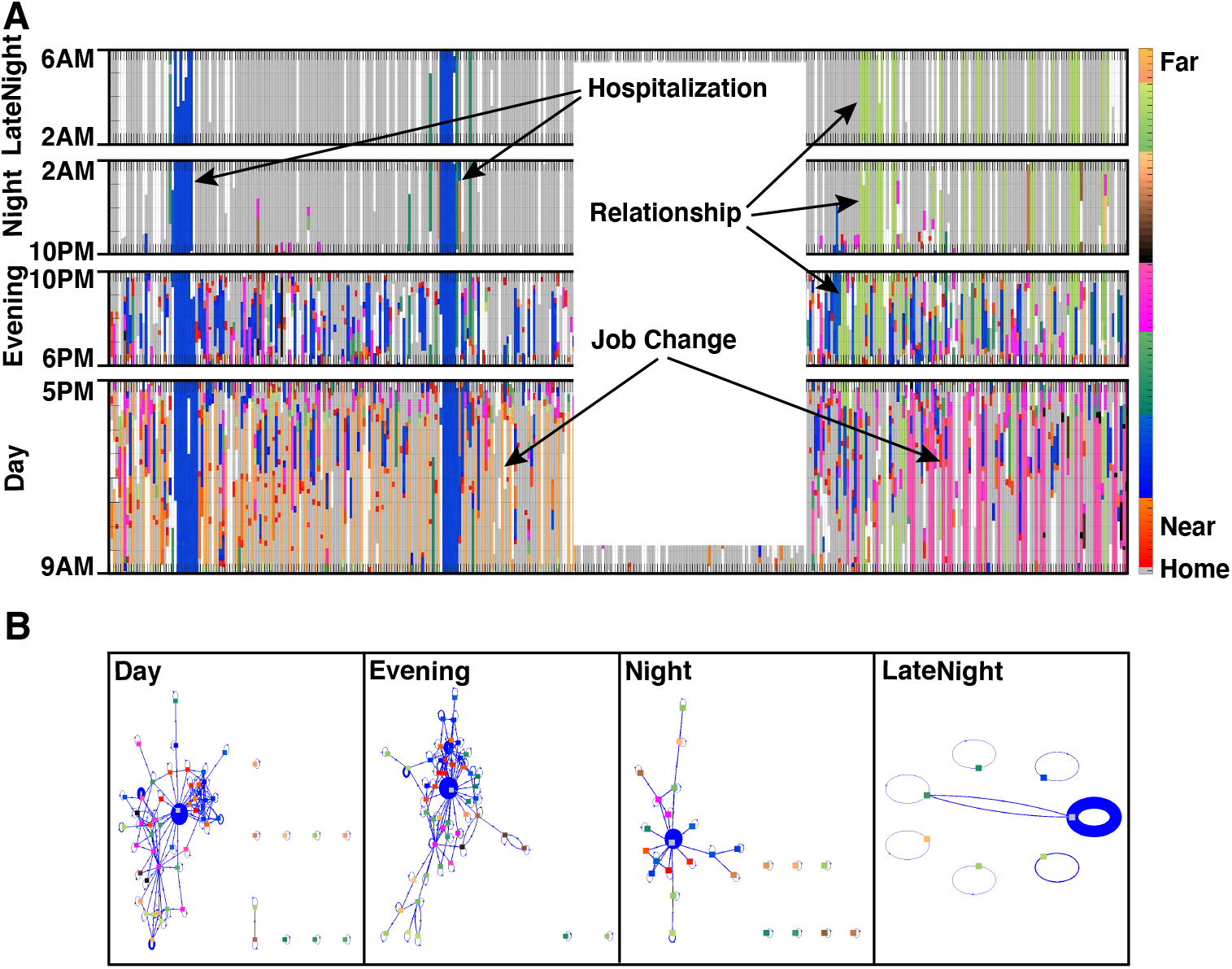
Detecting Major Life Events From Behavioral Time-Bands. The Behavioral Time-Bands **(A)** and Markov Model diagrams **(B)** are displayed for an individual diagnosed with a psychiatric disorder to illustrate the detection of major life events. The time intervals are modified from the original to avoid identifiability. The first prominent events are two occasions of staying at a specific location (blue) throughout all four Behavioral Time-Bands, which is coincident with their in-patient hospitalization. The individual visits the same location during the Day or Evening Time-Bands on the other days. Another major event occurs when the individual starts a relationship which is associated with the light green location. As labeled in the figure, the participant starts visiting the place during the Day and Evening Time-Bands, and then later during the Night and LateNight Time-Bands. Almost at the same time, the location that the participant spends time during the Day Time-Band also changes from the orange to pink location which is consistent with a job change. All these events were confirmed after detection with the individual during the later visits and reports.

Another event that takes place later during the recorded life of the same individual likely represents a pattern that can be interpreted as beginning and advancing a new relationship. The individual starts to stay overnight at a location shown in light green, where visits to the same location during the Evening and Night Time-Bands had been increasing for the prior two weeks. Almost at the same time, the orange location is visited frequently, and for about 7 hours during the Day Time-Band is switched with the pink POI, which might be a marker of work or daytime activity change as labeled in Figure 10A. While the definite reasons for these changes are unknown, the pipeline is able to capture these changes in patterned behavior.

### LateNight Time-Band Adjustment

The Time-Band defaults are chosen based on a typical assumed schedule. However, this assumption does not always lead to similar behavioral capture for all individuals. As an example, the Markov Model diagrams during the LateNight Time-Band for five participants (P1-P5) are shown in Figure 11. The default LateNight Time-Band is shifted up to 2 hours backward and forward. Although the default assumption is reasonable for most of the participants, the model for the late sleeping individual (P1) shows that if we shift forward the Time-Band by 2 hours, we can detect the time that the individual stays at Home for several hours (single-node Markov Model diagram) and might be associated with the sleep time of this individual. Moreover, this table shows the sensitivity of the model to the LateNight time assumption for each individual. As an example, P3 and P4 are not sensitive to adjusting the LateNight Time-Band and show similar patterns of two locations and multiple locations, respectively. The analysis on P2 shows that the Time-Band can be shifted up to 2 hours later but not earlier for this individual, while the LateNight Time-Band for P5 can be shifted earlier but not more than one hour later. This individual appears to start daily activities sometime between 7AM and 8AM.

**Figure 11.**
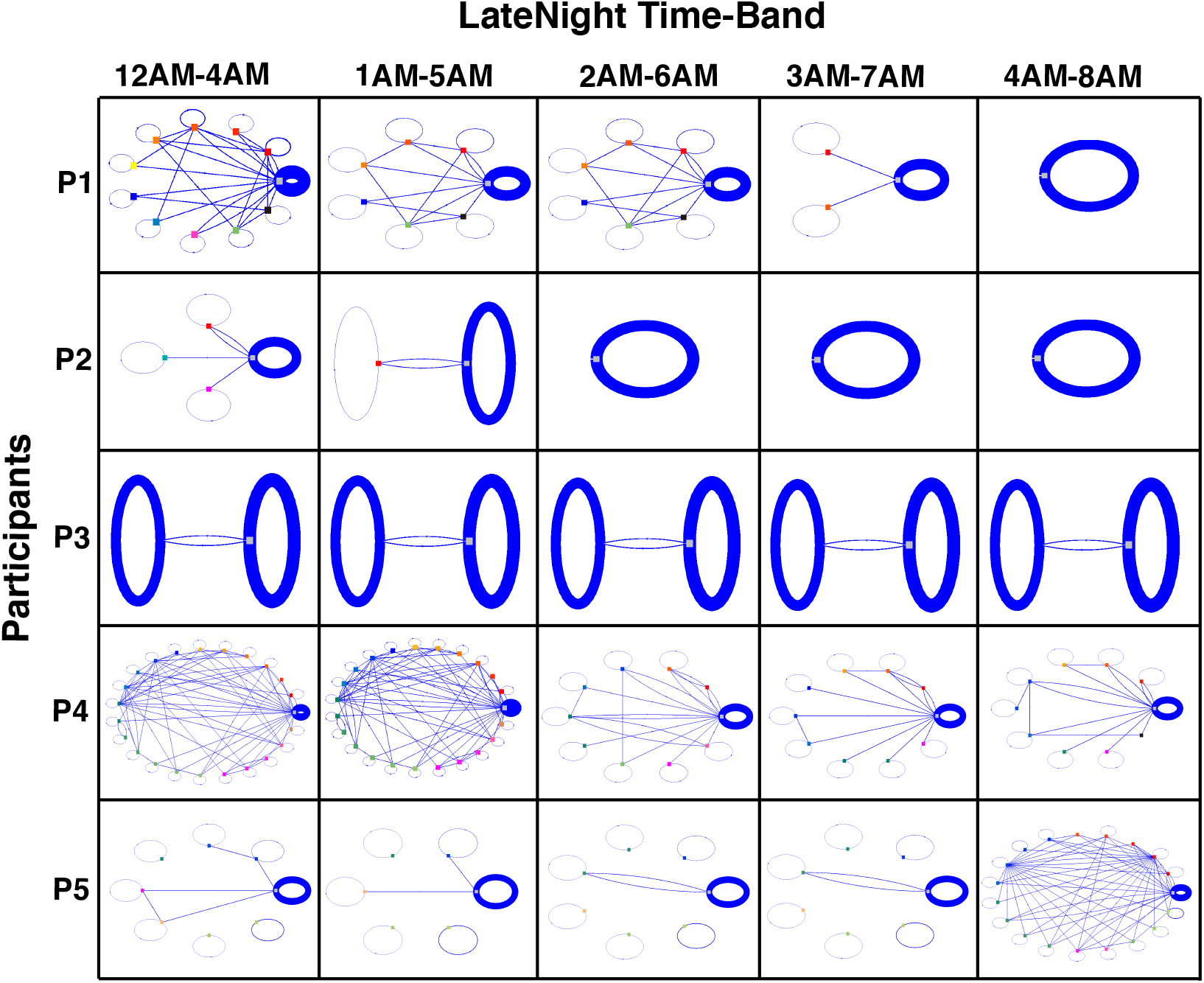
Effects of Temporally Shifting the LateNight Behavioral Time-Band. The Markov Model diagrams of the LateNight Time-Band for the previously presented participants are presented again as P1 to P5, respectively, with adjusting the default time of the LateNight Time-Band to begin from 12AM (leftmost column) to 4AM (rightmost column). The effect of temporal shifting of the Time-Band varies across the participants and is a reminder that analysis assumption can impact the resulting models. The default 2AM-6AM Time-Band bounds for LateNight appears a reasonable assumption but can capture residual evening activities (e.g., P1). Participants with markedly different sleep-wake cycles, such as shift workers, would likely be misunderstood without adjustment of the default Behavioral Time-Bands.

### Analysis of Weekly Behavioral Patterns

Another approach to increase the understanding of the individuals’ behavior based on location is to display the color-coded behavioral map separately for each day of the week with 24-hour Time-Bands. One example presented in Figure 12A shows the regular weekly pattern of a student’s (P6) life. For example, the orange location on Monday and Wednesday mornings likely indicates a two-day a week class or recurring activity, while dark blue could represent a more general location that could include classes, gym or dining hall, and the red location on the evenings of Wednesdays and Fridays might show a consistent socializing location or a different class. Another interesting observation from this analysis is the substantial difference between weekday and weekend patterns, consistent with prior deep-phenotyping analyses [18].

**Figure 12.**
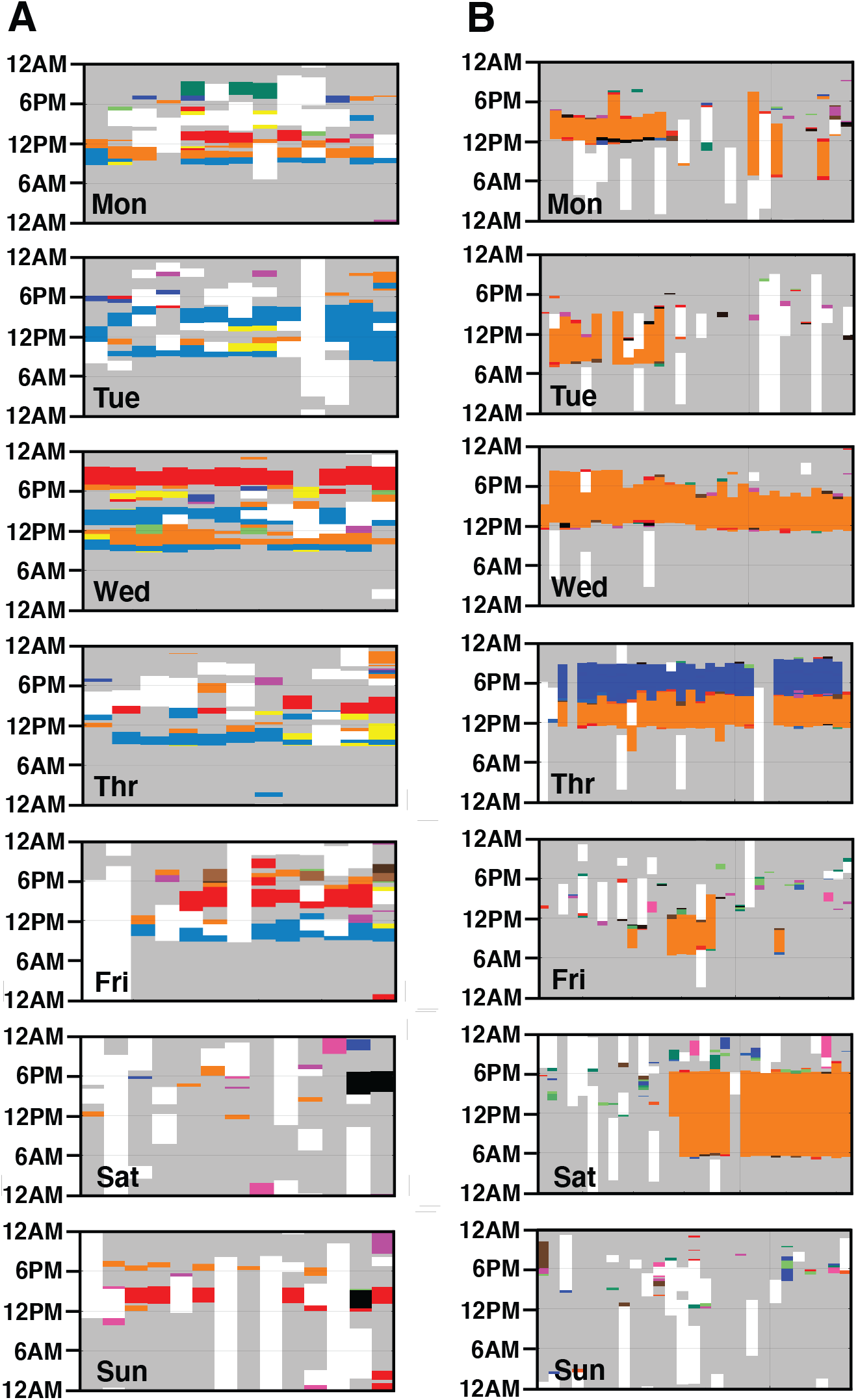
Locations Visited Show Informative Weekly Patterns. Maps are presented for separate days of the week to investigate the weekly schedules of an academic student **(A)** and an individual living with mental illness **(B)**. Each panel shows one day across many weeks to visualize structured patterns that are tied to the day of the week. The weekly schedule of the student (A) depicts regular visits to an orange location on Monday and Wednesday mornings and a red location on Wednesday evenings that likely represent classes or a recurring activity. The weekend days show irregular patterns. The weekly schedule of a participant living with severe mental illness reveals a distinct pattern (B). The individual rarely leaves Home except for going to a work location shown in orange. The orange (work) pattern reveals regular afternoon shifts on Wednesdays and Thursdays, and short shifts on Mondays and Tuesdays which are swapped by longer shifts on Saturdays midway through the sampling period. There is a regular Thursday evening location after work in dark blue that represents a recurring therapy session.

Another example is the weekly behavioral pattern of an individual living with severe mental illness (P7) who holds a part-time job (Figure 12B). The striking observation from the daily behavioral map of this individual, which is more revealing from the weekly pattern analysis, is that this participant barely leaves Home (gray) except for work (orange). The maps show regular afternoon shifts on Wednesdays and Thursdays, and long shifts on Saturdays, which are swapped later by short shifts on Mondays and Tuesdays.

### Mobility Patterns Versus Mood

Another co-product of DPLocate, in addition to the qualitative daily maps, is the ability to extract meaningful quantitative daily parameters related to mobility. Those parameters can be used in individuals to evaluate the effect of change in their daily routines as a clinical or life event measure. Some of those parameters presented here include (1) R (Kilometers): the radius of a circle that encompasses all Epochs visited in a day, (2) H (%): the percentage of the time the participant spends at the day’s dominant location which could be the Home POI if visited on that day or otherwise, the most visited place of the day, (3) P (#): number of POIs other than Home that the individual has visited during the day. As examples of their utility, we present the effect of shelter-in-place due to the Covid-19 pandemic on the mobility parameters as well as mood fluctuations of two individuals (P8, P9) living with psychiatric illness.

Figure 13 displays the results for P8. Even though this individual does not have a full-time job, there are multiple places (4-5 on average) other than Home that they visit often for more than an hour per day before the Covid-19 pandemic lockdown (day 172 for this participant). For the three months prior to the lockdown, the individual visits a location in red almost regularly every morning between 8AM to 9AM. The black line represents a moving average of the past 14-days and shows that an average of 80% of the time is spent staying at home. This routine is compromised after the lockdown, with more than 95% of the time spent at home and only 2-3 places visited per day each for brief periods.

**Figure 13.**
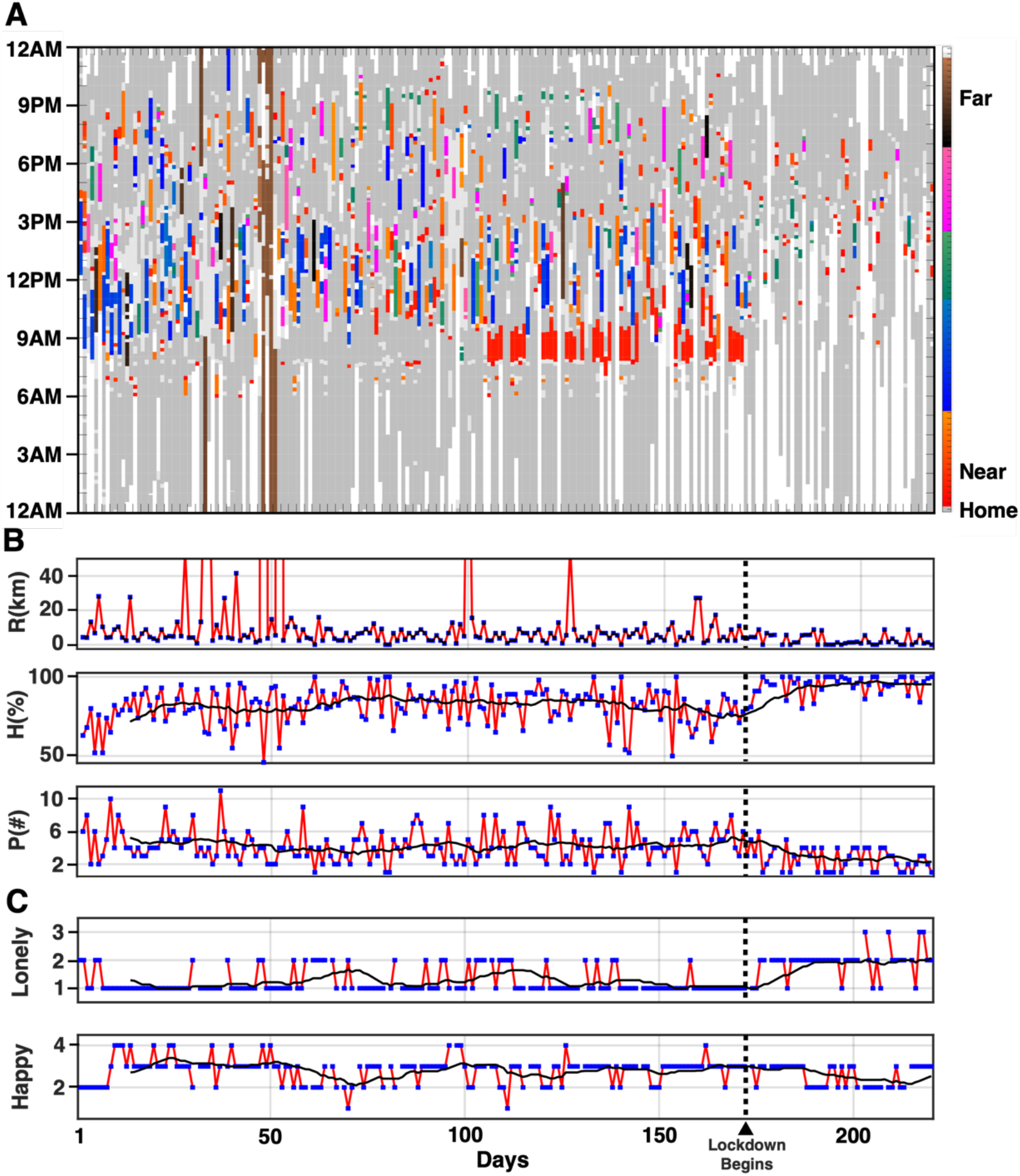
Mobility Pattern Reveals a Relation to Mood: Case 1. The Daily Map pattern **(A)** of a participant with severe mental illness is presented for 220 days along with extracted daily GPS parameters **(B)** and self-reported Happiness and Loneliness scores **(C)**. The daily GPS-extracted parameters are: (1) the radius of the circle that encompasses all visited Epochs during the day (R) in Kilometers, (2) the percentage of staying at home during the day (H) in percent, and (3) the number of POIs other than Home visited during the day (P). The 14-day backward moving average is depicted as black lines on H and P. (C) Daily self-reported Happiness and Loneliness are presented with a 14-day backward moving average depicted as black lines. The average trend shows that even though the individual does not spend a lot of time outside of their Home, the significant outside time reduces substantially around day 170 which coincides with the Covid19 pandemic lockdown. This change is associated with a marked increase in Loneliness and a less dramatic decrease in Happiness.

The effect of this routine change is paralleled by self-reports of increased Loneliness and decreased Happiness. These emotions are part of a larger daily self-report survey submitted through the Beiwe platform and reflect participant responses to daily questions of “Over the past 24 hours, how much were you feeling: Happy and Lonely” (separately). Each item was scored on a Likert scale of 0 (Not at all) to 4 (Extremely). The 14-day backward moving window reveals the increase in Loneliness and the decrease in Happiness, at least for the first two months after the onset of the lockdown.

Similar results are shown for P9, in Figure 14. This individual has a full-time job and lives in three places as home (gray, red, and brown) during the 1-year period of the study. The lockdown, which happened around day 320 for this participant, is associated with a significant drop in the number of places visited (P) and a significant increase in the percentage of time spent at home (H). This routine change is also associated with feelings of Loneliness.

**Figure 14.**
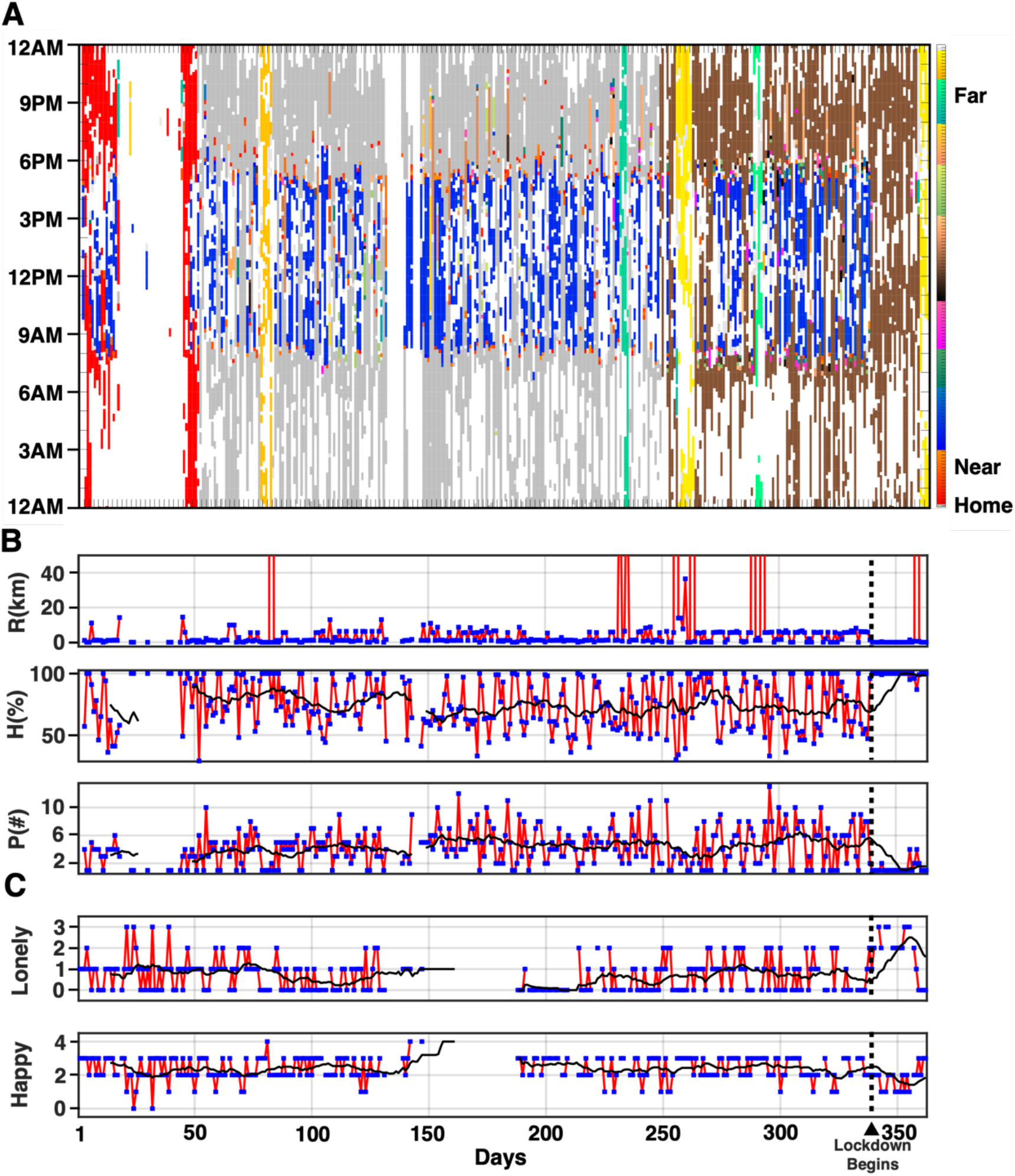
Mobility Pattern Reveals a Relation to Mood: Case 2. Similar to Figure 13, the Daily Map pattern **(A)**, extracted daily GPS parameters **(B)**, and self-reported Happiness and Loneliness scores are presented for 365 days for another participant with severe mental illness. This individual has a regular job, moves twice during the year without changing job locations and travels about five times. After day 335, the participant does not physically go to work due to the Covid19 pandemic lockdown and almost always stays at home. Self-reported Loneliness significantly increases, and Happiness decreases after the lockdown while mood is recovered when a travel event (shown in yellow) occurs after day 360.

## Discussion

The wide adoption of smartphones and wearable devices has led to growing interest in pipelines to extract meaningful features from GPS data. Here we introduce DPLocate, an open-source GPS analysis pipeline that securely analyzes encrypted raw GPS data. DPLocate uses temporal filtering to account for intermittency and artifacts caused by the inaccuracy and jittering of the GPS signal and then deploys a density-based spatial clustering algorithm to identify the most visited POIs. The patterning of visits to and from POIs, in turn, provides rich information about behavior, especially when analyzed within Time-Bands targeting distinct periods of the day. We discuss DPLocate in the context of the many evolving strategies to acquire and analyze GPS data.

Several prior studies collected GPS data by asking the participants to carry portable GPS devices, which are beneficial because such devices are optimized for continuous GPS tracking [4,30,34]. Many recent studies employ smartphones as the equipment to collect GPS data given their wide, existing utilization. The challenge of smartphones is that the GPS signal, in practice, is intermittent. To mitigate the effects of GPS signal gaps, studies have additionally exploited the Wifi access point [3] or beacon-based data mining [43] to increase the precision of the location detection and overcome the spontaneous missingness of the GPS signal. Other studies rely only on the signal provided by the GPS and deal with the data acquisition difficulties using filtering and classification algorithms [44,45]. The DPLocate pipeline takes such an approach by analyzing the raw GPS data using Epoch-based temporal filtering to remove artifacts and attenuate the effect of missingness.

### DPLocate’s Approach to Identifying Points of Interest

Multiple approaches have been put forward to analyze GPS data and extract relevant features based on the requirements and aims of different studies. Some applications focus on the mobility and activity of the participants with analysis of their trajectories [46], locomotion modes (walk, drive, ride), or direction of movement [40]. However, since the focus of the DPLocate pipeline is to investigate the behavior and life dynamic of individuals, it is aligned with studies that target individuals’ POIs [22] and rank them based on their significance [26], frequency of visit [31], or the length of time spent at the locations [47]. Clustering algorithms have been developed for the purpose of localizing GPS data that variably rely on temporal information (i.e., the locations where the person stays more or has a lower speed [31,48]), spatial information (i.e., the most visited locations [36,49]), semantic information (i.e., home, work, shopping and school categories [25]), or combinations of temporal, spatial and semantic information [7,27].

The DPLocate pipeline is built on spatial clustering after a temporal filtering process is employed to remove artifacts. Temporal clustering was not used to avoid bias that might result given a key output of DPLocate is the behavior of participants at different times of the day. Moreover, the pipeline was designed to be independent of semantic information (annotations of locations) to avoid reliance on proprietary semantic databases and potential security risks due to sharing the coordinates with the third party (although this can also be mitigated by storing local copies of coordinate reference databases). For spatial clustering, early efforts have deployed K-means [31], ordering index [50], and mixed distribution techniques [51]. DPLocate adopts an efficient recent strategy [24,27,35,37] by employing a revised version of DBSCAN, a density-based clustering algorithm, to find the POIs. Density-based algorithms benefit from some novel modifications such as dividing the space into specific grids or defining the minimum distance between clusters and are one of the most popular clustering algorithms for GPS data [49,52]. There are also other less utilized yet innovative approaches such as conditional random field [40] and kernel-based [44] algorithms that have great potential. For many studies, the analyses are complete after finding the significant POIs for the individual. However, this is the beginning point to model behavior and life dynamics.

### Behavioral Dynamics as a Phenotype

Insights into behavioral dynamics emerge when analyses investigate patterns of movements between POIs. As examples, studies have used POIs to measure the mobility and relocation of the individual during a day as an activity marker [53,54]. Accordingly, they extract various features such as location variance, entropy, homestay, total traveled distance, and circadian movement to quantify the GPS-related behavior of the individual during a day [1,39,55]. Another interest is predicting the next location an individual will visit based on the previous locations. Various versions of the Markov Model, such as the mobility Markov Model [37], mixed Markov Model [38], and n-step Hidden Markov Model [38] have been proposed and compared to alternatives such as hybrid dynamic mixed networks [41], tree-based hierarchical graphs [56], Bayesian Models [57] or other probabilistic models [42].

DPLocate specifically uses Markov Model to generate the diagram of transitioning between different locations at different Time-Bands for multiple detected phenotypes. The Markov Model diagrams in DPLocate are employed, as in Figure 11, to distinguish between phenotypes rather than predict the next location. The participants demonstrated clear differences in LateNight activities, as explicitly visualized in the Markov Model diagrams, which contained single location stays in some individuals (e.g., P2), dominant single locations with some variation (e.g., P4), and clear, patterned variation between two locations in others (e.g., P3).

### The Potential for Clinical Inference

A critical goal of behavioral studies that use GPS data is to make clinical inferences. Reported correlations between mobility markers extracted from GPS data and depression severity [1,19,55,58] and anxiety symptoms [5] make clear the potential utility of GPS data. Other studies have proposed GPS data mining to assess the mental health and academic performance of students [2,59] as well as individuals with bipolar [58,60,61] and schizophrenic [12,62,63] illness. Recent studies have offered candidate biomarkers of dementia and Alzheimer’s Disease in older adults using mobility patterns [64].

In the present work, patterns revealed through the DPLocate pipeline detected disrupted sleep as well as hospitalization in individuals living with psychiatric illness. Although investigating the association between the clinical symptoms and the location-related features is beyond the scope of this paper, the examples presented demonstrate the potential application of the pipeline to detect changes in behavior over extended periods of time, including examples where changes in GPS-derived measures of mobility associate with change in self-reported Loneliness and Happiness. Figure 13 displays a particularly clear example where dramatic changes in mobility occur when the Covid-19 pandemic lockdown begins.

### Caveats

The DPLocate pipeline provides a powerful tool to analyze intermittent GPS data and extract behavioral patterns of individuals based on their relocation. However, the approach has limitations. A common limitation of digital phenotyping tools, especially when analyzing GPS data, is sensitivity to missing data. GPS signal loss for extended periods during a day can cause the pipeline to miss visited locations and underestimate stay durations. However, DPLocate is by design robust to short term missingness (<12 minutes) which is more common in GPS data. Another limitation of the pipeline is its use of assumed Time-Bands to analyze the behavior of individuals. As was illustrated in Figure 11, the meaningful Time-Bands associated with daily routines such as sleep, work and socialization can vary across individuals. Dynamic Time-Band adjustment may be necessary to most accurately interpret the daily relocation patterns of some individuals.

## Data Availability

All data produced in the present study are available upon reasonable request to the authors

https://github.com/dptools/dplocate

## Acknowledgements

We thank Laura Farfel, Marisa Marotta, Erin Phlegar, Lauren DiNicola, Arpi Youssoufian, Elyssa Barrick, Nora Mueller, Crystal Blankenbaker, and Joanna Tao, for their help collecting data. Tim O’Keefe, Harris Hoke, Lily Jeong and Sarah Guthrie provided valuable assistance in neuroinformatics support. Kenzie W. Carlson helped with the Beiwe component of the studies. Work was funded by a generous gift from Kent and Liz Dauten, NIMH grants P50MH106435 and U01MH116925, and NIH award DP2MH103909.

